# Prevalence of dementia in elderly age population of Barangay Bangkal, City

**DOI:** 10.1101/2021.02.11.21251546

**Authors:** Vikash Jaiswal, Maria Kezia Lourdes Pormento, Namrata Hange, Neguemadji Ngardig Ngaba, Manoj Kumar Somagutta, Inna Celina Apostol Dy, Saloni Savani, Sana Javed, Shavy Nagpal, Arushee Bhatnagar, Mohit, Annie Singh, Dattatreya Mukherjee, Ruchir Paladiya, Freda Malanyaon

## Abstract

**Background:** Dementia, a significant cause of disability and dependency among older adults. The growing population of the elderly in the Philippines is expected to increase the prevalence of dementia in the country.

**Purpose:** This study aims to determine the prevalence of dementia in the elderly population of Barangay Bangkal, Makati City.

**Methods:** Descriptive cross-sectional community-based study was conducted to determine the prevalence of dementia in the elderly population of Barangay Bangkal, Makati City, aged 60 years and above over one month from mid-October to mid-November 2019. Data was collected with help of Mini-Mental State Examination – Philippines version (MMSE-P) to determine the cognitive status and diagnose dementia in elderly population.

**Results:** A total of 266 elderly adults participated in the study. Representatives of the study population were male (59.0%), married (68.0%), with an income of less than 5,000 peso (51.1%), working (64.3%), and with high school education (42.1%). The average age of the study population was 68.02 (± 6.76) years. The average MMSE score of the participants was 27.05 (± 3.94). The prevalence of dementia in the sample was 18.8%. Age, income, and level of education were associated with the MMSE score (*r*□ = - 0.26, *n* = 266, *p* < 0.001, *r*□ = 0.23, *n* = 266, *p* < 0.001, and *rs* = 0.19, *n* = 266, *p* = 0.002, respectively). The findings for statistical significance do resonate with clinical significance as evident during administration of MMSE score.

**Conclusion:** Advancing age increases the risk for cognitive decline while higher income and education level prevent or delays the onset of dementia. Collaborative management between the medical education faculty & students, researchers, and local state health officials might address dementia in the region.

## INTRODUCTION

### Background of the Study

Dementia, a significant cause of disability and dependency among older adults, requires urgent attention and care.^1-4^ It is a clinical diagnosis requiring new functional dependence based on progressive cognitive decline and represents a departure from previous mental functioning.^5^ Usually, dementia is of a chronic or progressive nature, characterized by deterioration in memory, thinking, behavior, and the capacity to make daily living (World Health Organization, 2019)^6^. Globally, around 50 million individuals have dementia with the majority of dementia cases reported from low or middle-income countries. This number is expected to rise to 82 and 152 million by 2030 and 2050 respectively. In 2015, it was estimated that dementia resulted in US$818 billion of global societal costs.^6,7,8^ In Asia-Pacific countries, including the Philippines, the cost was estimated at US$185 billion for 2015.^9^

Furthermore, dementia has detrimental effects on caregivers and families.^10-12^ Dementia grossly affects the physical, mental, and social aspects of health and health quality, contributing to a higher economic burden. ^13-16^. Researchers have postulated that dementia is often accompanied by emotional control, social behavior, and motivation to thrive in life. It also puts the elderly at risk for malnutrition.^17-19,20-21^ Orsitto et al, conducted a study on 623 elderly patients who were hospitalized for mild cognitive impairment, and concluded 58% risk of malnutrition while 24% were malnourished ^22^.

### Diagnosing dementia

A qualitative study revealed that many South Asians perceive dementia as a part of normal aging process, and thus are unlikely to believe that dementia could be treated.^23^ However, previous studies had identified genes, depression, bereavement, brain injury, and stressful life as contributing factors to the disorder.^24-25^Keeping in consideration the different types of dementia (Alzheimer’s, Vascular, frontotemporal, and dementia with Lewy bodies), physicians face challenges in terms of differentiation, and management.^26^ The most important dementia causes were identified as Alzheimer’s dementia (85.5%) followed by vascular dementia (11.7%) in the Philippines during a study by Dominguez et al.^24-25^ A few reasons quoted for difficulty diagnosing dementia were lack of precise screening tools, inadequate and insufficient time for patient consultation required for identification of symptoms and diagnosis.^27,28^ Globally, a widely used screening tool for dementia is the Mini-Mental Status Exam (MMSE).^29^ However, the MMSE test can be difficult to administer because patients memory and cognitive function are erratic, fluctuating, and unpredictable in dementia cases.^30-32^ In Dementia assessment, the patient might give vague answers reflecting mental stability while, in reality, patients are struggling and attempting to cover up problems. Another flaw of this assessment tool is the staff’s tendency to give subtle hints of the correct answer to patients subconsciously, and such shortcomings make MMSE far from an error-proof tool. Thereby, we may not be able to diagnose dementia solely based on the numbers provided from the test (Coin et al., 2012).^33-34^

### Purpose of study

Barangay Bangkal is a barangay (village) in Makati City located in Metro Manila, Philippines where study was conducted due to convenience of location, and considering the tight deadline of thesis/project. This study outlines the prevalence of dementia in Barangay Bangkal, Makati City, and its association with socio-demographic factors.

An estimated 5-8% of the elderly in The Philippines have dementia defined as individuals aged 60 years of age and older.^35,36,37^ Due to advances in medical modalities leading to longevity and increased elderly population, a growing trend of dementia is also expected in the Philippines. The Geriatrics Infolink Management Strategies Philippines Inc.(2018) has reported around 301,000 Filipinos with dementia by 2015. This organization had predicted that these cases were expected to reach upto 568,000 and 1.15 million by 2030, and 2050 respectively.^36-37^ This upward trend along with the increasing population of the elderly in the Philippines ^24^ calls for studies that will look further into the prevalence and pattern of dementia in the country. Considering the negative impacts of dementia on the individual, caregivers, families, and society, studies that examine dementia in the Philippines remain scarce.

## Methods

### Research Design and Sampling Technique

This is a descriptive cross-sectional community-based study conducted to determine the prevalence of dementia in the elderly population of Barangay Bangkal, Makati City. This study was conducted over one month from mid-October to mid-November 2019 and included elderly residents of at least 60 years. The target population was composed of 4,500 elderly adults, identified through the barangay census. The researchers divided the barangay into east and west sections. Streets were chosen at random, whereby house to house visit was done to gather respondents. Individuals who fit the criteria and were willing to participate were given a printed consent form, and the interview was conducted on the same day. The estimated target sample size was 355, which was calculated by power analysis. The power was set at 80%, confidence level at 95%, and margin of error at 5% (SurveyMonkey, 2019).

### Exclusion Criteria

Elderly adults with an acute severe health crisis, end stage disease or terminal illness, severe visual and hearing impairment, previous psychiatric condition, and inability to understand the consent process were excluded from the study.

### Research Instrument

The study used the Mini-Mental State Examination – Philippines version (MMSE-P), a validated translation of the original screening test to determine the cognitive status and diagnose dementia in the elderly population.^38-39^ A copy of the MMSE-P is attached in Appendix A. The MMSE-P measured the cognitive impairment of older adults by examining functions including registration, attention, calculation, memory, language, and orientation, and scores were ranged from 0 to 30 points. MMSE scores were considered in the range of 25 - 30 where 20 - 24 was mild, 13 - 20 moderate, and < 12 severe dementia..13

### Ethical Consideration

The study was abided by the American Medical Association School of Medicine’s ethical guidelines in conducting research. Participants were briefed on the objective of the study, and written informed consent was obtained. All participants signed the informed consent to confirm participation, and were assured about the privacy and confidentiality of data.

### Data confidentiality

Only the researchers had access to the collected data which was encoded in MS Excel in a password protected laptop, and kept in a secured locked room. The MMSE-P response sheet was shredded and discarded a month after the completion of the study which was in accordance with respecting the patient’s wishes. No personal identifiers were included in the analysis or published in the report.

### Data Collection

Data is collected with the MMSE-P, a 30-item questionnaire used to measure the cognitive impairment in an individual and is used to screen for dementia. It is also used to measure the level of cognitive impairment, and examines functions, including registration, attention, calculation, memory, language, and orientation. The researchers followed the recommended script, timing, and scoring as specified for the MMSE-P to ensure the examination’s sensitivity and specificity. Before the assessment, the instruction guide, MMSE-P response sheet, a pencil, a wristwatch, the command card for the “close your eyes” reading comprehension subtest, the template for pentagon figure copying and blank sheets of paper were prepared by the researchers. The verbal instructions were given in Tagalog (native language) or in English based on the participants’ preference. The instruction sheet was not shared with the subject during the test administration. At the end of the test, the total score was tallied. The Research team was briefed and helped in the administration of MMSE, and verbal and written consent was obtained prior to testing.

### Variables

The Independent variables were demographic data, including age, sex, marital status, income, occupation, nationality, and education level. The outcome of interest was the level of impairment of cognition in dementia.

### Statistical Analysis

Study participant’s characteristics, the prevalence of dementia, and impairment levels related to dementia were analyzed using descriptive statistics. Continuous data were presented as mean and standard deviation, while categorical variables as number and percentages. Pearson’s correlation coefficient, chi-square test of independence/association, or Spearman correlation was done to assess the relationship between MMSE scores and demographic variables. Results were considered statistically significant if *p* < 0.05. Missing data were included in the analysis, and SPSS version 22 was used to analyze the data.

## RESULTS

### 1. Socio-Demographic Profile of study participants

A population-based cross-sectional study was conducted among a total of 266 participants from mid-Oct to mid-Nov 2019 to evaluate the prevalence of dementia and related impairments in the elderly population of Barangay Bangkal, Makati City. Table 1 as given below, summarizes the socio-demographics of the study participants. The mean age of participants was 68.02 (+ 6.76) years. The gender distribution was 59% male and 41% female representing a sex ratio of 1.44. Most of them were male (59.0%), married (68.0%), with an income of less than L5,000 (51.1%), leading active work life (64.3%), and with high school education (42.1%).

**Table 1.** Summary of Participants’ Demographics Data

| Parameter | *n* (%) |
| --- | --- |
| Age | 68.02 (6.76)* |
| Sex |  |
| Male | 157 (59.0) |
| Female | 109 (41.0) |
| Marital status |  |
| Married | 181 (68.0) |
| Single | 62 (23.3) |
| Others | 23 (8.6) |
| Income |  |
| Less than ₱5,000 | 136 (51.1) |
| ₱5,000 – ₱9,999 | 76 (28.6) |
| ₱10,000 and above | 54 (20.3) |
| Occupation |  |
| Working | 171 (64.3) |
| Retired | 95 (35.7) |
| Level of education |  |
| High school | 112 (42.1) |
| College | 76 (28.6) |
| Elementary | 38 (14.3) |
| Vocation | 21 (7.9) |
| No education | 11 (4.1) |
| Post-graduate | 8 (3.0) |

\* Age is presented as mean (standard deviation)

### 1. Prevalence of dementia and the level of impairment

Table 2 presents the participants’ MMSE score and level of impairment. The mean MMSE score of the study participants was 27.05 (± 3.94). Majority of patients have normal cognition (216, 81.2%). Out of the 266 participants, 50 (18.8%) of them have dementia with various severity on MMSE assessment. Out of 50 dementia patients, 30 (11.3%) had mild dementia, 16 (6.0%) had moderate dementia, and 4 (1.5%) had severe dementia.

**Table 2.** Summary of the Participants’ MMSE Scores and Level of Impairment

| Parameter | n (%) |
| --- | --- |
| MMSE score | 27.05 (3/94)* |
| Level of impairment |  |
| Normal | 216 (81.2) |
| Mild dementia | 30 (11.3) |
| Moderate dementia | 16 (6.0) |
| Severe dementia | 4 (1.5) |

\* mean (standard deviation)

### 2. Association between MMSE score and demographic variables

Table 3 shows the association between MMSE scores, age, income, and level of education. The data for age was non-linear, with outliers, and non-monotonic; thus, Kendall’s tau-b (τb) correlation coefficient was done. There was a weak negative correlation between MMSE score and age, *r*_□_ = −0.26, *p* < 0.001. Similarly, the data for income was monotonic, hence, Kendall’s tau-b (τb) correlation coefficient was used. The analysis revealed a weak positive correlation between MMSE score and income, *r*_□_ = 0.23, *p* < 0.001. On the other hand, a Spearman correlation was computed to assess the relationship between MMSE score and level of education. There was a weak positive correlation between the variables, *rs* = 0.19, *p* = 0.002.

**Table 3.** Results of Correlation Between Demographic Variables and MMSE Scores

| Variable | MMSE Scores |
| --- | --- |
| Age | -0.26** |
| Income | 0.23** |
| Level of education | 0.19* |

$p < 0.05^*$ , $p < .01^{**}$

Table 4 as below summarizes the association between level of impairment and sex, marital status, occupation using the Chi-square test of independence/association. The analysis revealed a non-significant interaction between level of impairment and sex, *χ*2(3) = 1.46, *p* = 0.69. Men were not as likely to get dementia than women. Similarly, there was no significant association between the level of impairment and marital status, *χ*2(6) = 10.80, *p* = 0.10. Thus, single individuals were not more likely to suffer from dementia than married individuals or those with another marital status. There is also a non-significant correlation between level of impairment and occupation, *χ*2(3) = 2.90, *p* = 0.41. Working individuals were not more likely to suffer from dementia than retirees.

**Table 4.** Results of Correlation Between Demographic Variables and Level of Impairment

| Variable | Level of Impairment |
| --- | --- |
| Sex | 0.69 |
| Marital status | 0.10 |
| Occupation | 0.41 |

$p < 0.05^*$ , $p < .01^{**}$

## DISCUSSION

This population-based cross-sectional study was undertaken mainly to determine the prevalence of dementia in the elderly population of Barangay Bangkal Makati City. This study reported a prevalence of 18.8% for dementia, with the majority for mild dementia (11.3%), followed by moderate (6.0%) and significantly less proportion of severe dementia (1.5%). Dementia prevalence in this study is higher than the average global incidence of dementia at 5-8%.^2,7,24,35^ This prevalence has exceeded the prevalence reported in a study conducted in Marikina City of the Philippines, which was 10.6%.^24^Nevertheless, it was lower than the prevalence (32.2%) obtained by Abat, Reyes, and Ramos (2009) in their study of elderly Filipinos^.40^ This study had its limitations in collecting data with purposive sampling. Considering that the current study could not meet its target sample size, the inclusion of additional data might have influenced the prevalence of dementia.

This study has reported age to have a weak negative correlation with the MMSE scores, *r□* = −0.26, *p* < 0.001. Thus, advancing age has been linked with lower MMSE scores and possible dementia in the current study. This is comparable to the results of previous studies reporting the increasing risk of dementia with advancing age.^41,42^ Furthermore, prior studies found that incidence and prevalence of dementia accelerate after 65 years and double at around 5-6 years until the age of 90.^43,44^This current study findings are crucial for public health, considering the growing population of older adults in the Philippines. Badana and Andel (2018) claimed that the population of Filipinos ages 60 to 79 years old is expected to grow by 4.2% while those who are 80 years and older are expected to increase by 0.4% from 2010 to 2030.^45^

The current study data also showed income to have a weak positive correlation with MMSE scores and income, *r□* = 0.23, *n* = 266, *p* < 0.001. This implies the protective effect of income on cognitive decline. A comparable result was obtained by Scazufca et al. (2008), who found that elderly individuals with low income were at greater risk for dementia.^41^ They attributed this to the failure of this group to reduce morbidity at old age. Also, Brayne et al. (2006) found that improved social class can prevent the risk of cognitive decline.^46^ The discrepancy brought about by better socioeconomic status may be attributed to differences in preventable health- and lifestyle-related factors.^47^Elderly persons with higher income tend to have better access to a healthier lifestyle than those in the lower socioeconomic status. Therefore, the former has a more significant opportunity of reducing their risk of dementia.

A higher level of education can also prevent dementia, while the results obtained from the current study found a weak positive correlation between the level of education and MMSE scores, rs = 0.19, n = 266, p = 0.002. Similar findings were reported by studies done in Jalisco, Mexico and São Paulo, Brazil.^41,42^ These prior research revealed that individuals with a higher education level are at a reduced risk for developing dementia.^41,42^ Nevertheless, a systematic review done by Sharp and Gatz (2012) argued that the years of education could only attenuate the risk for dementia if schooling corresponded to greater learning and better cognitive stimulation.^48^ This is aligned with the theory of cognitive reserve, which claims that individuals who experienced mental and physical stimulation through education had better cognitive functioning, which later acts as a protective mechanism against dementia.^49^

The current study found no gender differences in the incidence of dementia, *χ*2(3) = 1.46, *p* = 0.69. This contradicts the study written by Velazquez-Brizuela et al. (2014), who reported that dementia was more common in women than men.^42^ The results from the Swedish Twin Registry aged 65 years and above also found that dementia was greater in women after the age of 85 years.^50^ The higher incidence among females is attributed to various factors. This includes longevity, biology, cognition, and social roles and opportunities.^51^ One reason that the current study yielded contradicting results could be the fact that the study participants involved were relatively younger than those involved in prior studies (*M* = 68.02 + 6.76 years vs. 71.6 ± 8.30 years for Velazquez-Brizuela et al. 2015 ^42^ and 72.01 + 8.94 years for Beam et al., 2018.^50^ No subgroup analysis was also done in the current study to determine if women after 85 were at greater risk.

Marital status was not associated with level of impairment, *χ*2(6) = 10.80, *p* = 0.10. Thus, being single does not increase the risk of developing dementia. This contradicts the findings of Velazquez-Brizuela et al. (2014)^42^ and Sundstrom, Westerlund, and Kotrylo (2015).^52^ These prior studies claimed that social relationships could reduce the risk of dementia. Other factors could have influenced the result of this current study. For instance, in the Philippines, families are closely knit, and it is common for elderly Filipinos to live with relatives. Thus, even though some participants are not married, it is likely that they are not living alone.

The results also found that occupation is not associated with the level of impairment, *χ*2(3) = 2.90, *p* = 0.41. In other words, the risk of developing dementia among those who work and are retired are similar. This opposes the results from a study done by Dufouil et al. (2014),^53^ who found that per extra age at retirement reduces the risk of dementia by 0.968 (95% CI [0.96, 0.97]). They inferred that working at an old age is linked with better levels of cognitive and social stimulation. Nevertheless, Ihle et al. (2016) argued that earlier retirement does not necessarily result in cognitive decline.^54^ They found that if elderly individuals are involved in a moderate number of leisure activities, they are likely to prevent or delay dementia. Engaging in leisure activities at old age may provide cognitive enrichment.

This is the first study to examine the prevalence of dementia in the elderly age population of Barangay Bangkal, Makati City. Thus, the results can provide the basis for policy development in the community, public health interventions, and further research to address the dementia epidemic—the high prevalence rate of dementia warrants immediate action to further research and public health management. Collaborative management from future students and barangay officials is necessary for addressing dementia in the site.

Overall, the literature synthesis illuminates the need for policy development and research for dementia that will address the needs of individuals who will continue to suffer from dementia. This current study will be created for the sake of addressing this identified knowledge gap. By identifying the prevalence of dementia in Barangay Bangkal, Makati City, this study aims to address more information regarding the current dementia trend in the Philippines.

### Limitations of the study

This study is a baseline evaluation of the resident’s data and no predictors were presented. The sample size obtained was relatively small compared to the target sample size of 355 elderly adults due to time constraints and some potential participants’ refusal to join the study. Due to the time constraint and refusal of some potential participants to participate, the targeted sample size was not met. Moreover, the prevalence of dementia was only an estimate. Specialists should confirm the diagnosis of dementia.

### Conclusion

The prevalence of dementia in the elderly age population of Barangay Bangkal, Makati City, is relatively high at 18.8%, with the majority reporting mild and moderate dementia. Advancing age is linked with increased risk for cognitive decline, while higher income and education levels have preventive dementia effects. The baseline data obtained in the current study can be used to design health policies in the barangay. However, these results should be interpreted with caution because of the relatively small sample size.

## Data Availability

All the details are mentioned in the Method section of the study.

## Abbreviation

MMSE-P: Mini-Mental State Examination – Philippines version
MMSE: Mini-Mental State Examination

## Acknowledgement

We thank all the patients who participated in the study, and families for their support.

## Ethical Statement

This study was approved by the institutional ethical committee of AMA School of Medicine, Makati, Philippines.

## Author Contributions

Dr. Jaiswal - Conceived the main idea, study design including data collection, conceptualization, interpretation, and workflow along with Dr. Mohit et al.

Dr. Pormento, Dr. Hange, Dr. Ngaba, Dr. Somagutta, Dr. Nagpal worked on conceptualization, review, data analysis, and editing of the study

Dr. Apostol Dy helped in Research design, Drafting, and approval of the version for final publishing.

Dr. Javed, Dr. Singh, Dr Bhatnagar, Dr Savani, Dr Paladiya and Dr. Malanyaon Dr Dattatreya helped in Reviewing the study,

